# High Seroconversion Rates Amongst Black and Hispanics With Hematologic Malignancies after SARS-CoV-2 Vaccination

**DOI:** 10.1101/2021.09.13.21263365

**Authors:** Lauren C Shapiro, Astha Thakkar, Radhika Gali, Jesus D Gonzalez-Lugo, Abdul-Hamid Bazarbachi, Shafia Rahman, Kith Pradhan, Karen Fehn, Michelly Abreu, Noah Kornblum, Kira Gritsman, Mendel Goldfinger, Aditi Shastri, Ioannis Mantzaris, Ira Braunschweig, Balazs Halmos, Amit Verma, Margaret McCort, Lizamarie Bachier-Rodriguez, R. Alejandro Sica

**Author notes:** **Corresponding Author:** R. Alejandro Sica, MD, Department of Oncology, Montefiore Medical Center, Albert Einstein College of Medicine, 111 East 210th Street, Bronx, NY 10467.

## Abstract

It is well established that COVID-19 carries a higher risk of morbidity and mortality in patients with hematologic malignancies, however, very little data on ethnicity specific responses in this particular patient population currently exist. We established a program of rapid vaccination and evaluation of antibody-mediated response to all EUA COVID-19 vaccines in an inner city minority population to determine the factors that contribute to the poor seroconversion to COVID-19 vaccination in this population. We conducted a cross-sectional cohort study of 126 patients with hematologic malignancies in the outpatient practices of our institution who completed their vaccination series with one of the three FDA EUA COVID-19 vaccines, Moderna, Pfizer, or Johnson & Johnson (J&J). We qualitatively measured Spike IgG production in all patients using the AdviseDx SARS-CoV-2 IgG II assay and quantitatively in 106 patients who completed their vaccination series with at least 14 days after the 2^nd^ dose of the Moderna or Pfizer vaccines or 28d after the single J&J vaccine. Patient characteristics were analyzed using standard descriptive statistics and associations between patient characteristics, cancer subtypes, treatments, and vaccine response were assessed using Fisher Exact test or Kruskal-Wallis Rank Sum test. The majority of patients (74%) were minorities. Seventy patients (60%) received Pfizer, 36 patients (31%) Moderna, and 10 patients (9%) J&J. We observed a high-rate of seropositivity (86%) with 16 pts (14%) having a negative Spike IgG. Of the 86 minority patients included, 94% Blacks (30/32) and 87% (39/45) Hispanics showed seropositivity. The factors that contributed to significantly lower seroconversion rates included patients with Non-Hodgkin lymphoma (p=0.005), those who received cytotoxic chemotherapy (p=0.002), IVIG (p=0.01), CAR-T cell therapy (p=0.00002), and CD20 monoclonal antibodies (Ab) (p=0.0000008). Plasma cell neoplasms (p=0.02), immunomodulatory agents (p=0.01), and proteasome inhibitors (p=0.01) had significantly higher seroconversion rates, and those with a history of prior COVID-19 (11%, 12/106) had significantly higher antibody titers (p=0.0003). The positivity rate was 86% (37 seropositive, 6 seronegative) for autologous HSCT and 75% (3 seropositive, 1 seronegative) for allogeneic HSCT. No life-threatening AE were observed. We show high seroconversion rates after SARS-CoV-2 vaccination in non-White patients with hematologic malignancies treated with a wide spectrum of therapeutic modalities. Vaccination is safe, effective, and should be encouraged in most patients with hematologic malignancies. Our minorities based study could be employed as an educational tool to dispel myths and provide data driven evidence to overcome vaccine hesitancy.

---

It is well established that COVID-19 carries a higher risk of morbidity and mortality in patients with hematologic malignancies.^1,2^ The mRNA-based vaccines BNT162b2 (Pfizer) and mRNA-1273 (Moderna), and adenovirus-based Ad26.COV2.S (Johnson & Johnson) vaccine have robust safety and efficacy against COVID-19 among immunocompetent individuals, however, patients with cancer were not enrolled in these registration trials.^3-5^ Emerging evidence suggests that despite the three COVID-19 vaccines with emergency use authorization (EUA) by the FDA inducing high levels of immunity in the general population, patients with hematologic malignancies have lower rates of seroconversion for the SARS-CoV-2 Spike IgG antibody (Spike IgG) and thus possibly lower protection against severe COVID-19.^6-8^ In particular, smaller subgroups of patients with certain hematologic malignancies including chronic lymphocytic leukemia (CLL), multiple myeloma, and patients treated with B-cell depleting therapies (i.e. anti-CD20 monoclonal antibodies (MoAb), chimeric antigen receptor T-cell [CAR-T] therapy) and hematopoietic stem cell transplantation (HSCT), have shown markedly impaired antibody-mediated responses to COVID-19 vaccines.^9-12^ There currently exists, however, very little data on ethnicity specific responses in this particular patient population. We established a program of rapid vaccination and evaluation of antibody-mediated response to all EUA COVID-19 vaccines in an inner city minority population to determine the factors that contribute to the poor seroconversion to COVID-19 vaccination in a broad range of patients with hematologic malignancies.

We conducted a cross-sectional cohort study of patients with hematologic malignancies in the outpatient practices of our institution between March 29, 2021 and July 8, 2021. Participants were enrolled after signing informed consent if they received one of the three FDA EUA COVID-19 vaccines, Moderna, Pfizer, or Johnson & Johnson (J&J). We qualitatively and quantitatively measured Spike IgG production using the AdviseDx SARS-CoV-2 IgG II assay designed to detect IgG antibodies directed against the receptor-binding domain of the S1 subunit of the SARS-CoV-2 spike protein. This assay has shown high sensitivity (95.6%) and 100% positive percent agreement with other platforms in both the post-COVID-19 infection and post-vaccination settings.^13^ Quantitative analysis was performed on patients who completed their vaccination series at least 14 days after the second dose of the mRNA-based vaccines, or 28 days after the single J&J vaccine. Patient characteristics were analyzed using standard descriptive statistics and associations between patient characteristics, cancer subtypes, treatments, and vaccine response were assessed using Fisher Exact test (categorical variables) or Kruskal-Wallis Rank Sum test (categorical and ordinal variables). Statistical significance was determined at α<0.05. All analyses were performed in R (version 3.6.2). Study protocol, data collection, and analysis was approved by Montefiore Medical Center Institutional Review Board.

A total of 121 patients were enrolled by informed consent and another 10 patients included by retrospective chart review. Five patients did not have Spike IgG performed after consent and excluded. Ten patients had Spike IgG testing before vaccination series completion and excluded from quantitative analysis. A total of 116 patients were included in immunogenicity analysis and 106 patients in quantitative analysis. Baseline characteristics and representative hematologic malignancies are listed in Table 1. The majority of patients (74%) were minorities. Seventy patients (60%) received Pfizer, 36 patients (31%) Moderna, and 10 patients (9%) J&J. Median time from vaccination completion to Spike IgG was 40 days. We observed a high-rate of seropositivity, 86% (100 patients) with 14% (6 patients) having a negative Spike IgG. Of the 86 minority patients included, 94% Blacks (30/32) and 87% (39/45) Hispanics showed seropositivity. Median Spike IgG titer was 2157 AU/mL (1697 AU/mL, 5290 AU/mL, and 1078 AU/mL for Pfizer, Moderna, and J&J respectively), although percent positivity and titer was not statistically significant between vaccine types. We observed significantly lower seroconversion rates (70%) in patients with lymphoid malignancies, specifically Non-Hodgkin lymphoma (p=0.005); however, the seropositive rate was 96% and 98% in patients with myeloid and plasma cell neoplasms respectively. Patients who received cytotoxic chemotherapy (p=0.002), IVIG (p=0.01), CAR-T therapy (p=0.00002), and anti-CD20 MoAb (p=0.0000007), especially within 6 months of Spike IgG evaluation (p=0.02), also showed significantly lower seroconversion rates. Use of BCL2 inhibitors (p=0.04), anti-CD20 MoAb (p=0.0009), CAR-T therapy (p=0.01), BTK inhibitors (p=0.04), current steroid use (p=0.002), and IVIG (p=0.003) also correlated with significantly lower antibody titers with a trend toward lower antibody titers in patients on any active cancer therapy at time of vaccination (p=0.051). Plasma cell neoplasms (p=0.02), immunomodulatory agents (p=0.01), and proteasome inhibitors (p=0.01) had significantly higher seroconversion rates, and patients with history prior COVID-19 (11%, 12/106) had significantly higher antibody titers (p=0.0003). Of 47 patients who received HSCT, 43 received an autologous HSCT and 4 an allogeneic HSCT. Median time to HSCT was 30.1 months. The positivity rate was 86% (37 seropositive, 6 seronegative) for autologous HSCT and 75% (3 seropositive, 1 seronegative) for allogeneic HSCT with no significant association with seroconversion, antibody titer, or time (greater or less than 1 year) since transplant. All patients who received anti-CD19 (Axi-cel) CAR-T therapy (0/6) were seronegative, and 1 patient that received BCMA-directed CAR-T therapy (Cilta-cel) was seropositive with no association between timing CAR-T cell infusion (greater or less than 1 year) and seroconversion or titer (median time to CAR-T therapy 17.5 months). The majority of patients, 64% and 53%, reported no adverse effects (AE) to the 1^st^ and 2^nd^ dose respectively. The most common AE were mild in severity and included sore arm, muscle aches, fatigue, and fever. No life-threatening AE were observed.

**Table 1.** Baseline Characteristics and Types of Cancer Therapy.

|  | **Total**<br>**(N=116)** | **Spike**<br>**Antibody**<br>**Positive**<br>**(N=100)** | **Spike**<br>**Antibody**<br>**Negative**<br>**(N=16)** | **P-value** |
| --- | --- | --- | --- | --- |
| **Patient Characteristics** |  |  |  |  |
| Age (years), median [range] | 70.5 [27-91] | 70.5 [27-89] | 69 [32-91] | 0.11 |
| Sex (male), n (%) | 65 (56%) | 53 (53%) | 12 (75%) | 0.11 |
| Race, n (%) |  |  |  | 0.36 |
| Black | 32 (28%) | 30 (30%) | 2 (13%) | 0.23 |
| Hispanic | 45 (39%) | 39 (39%) | 6 (38%) | 1.00 |
| White | 30 (26%) | 23 (23%) | 7 (44%) | 0.12 |
| Asian | 2 (1%) | 2 (2%) | 0 (0%) | 1.00 |
| Other/Unknown | 7 (6%) | 6 (6%) | 1 (6%) | 1.00 |
| Hematologic malignancy, n (%)* |  |  |  | 0.003** |
| Acute Leukemia | 8 (7%) | 8 (8%) | 0 (0%) | 0.60 |
| Castleman Disease | 1 (1%) | 1 (1%) | 0 (0%) | 1.00 |
| Hodgkin Lymphoma | 4 (3%) | 3 (3%) | 1 (6%) | 0.45 |
| MDS | 18 (16%) | 17 (17%) | 1 (6%) | 0.46 |
| MPN | 3 (3%) | 3 (3%) | 0 (0%) | 1.00 |
| Non-Hodgkin Lymphoma | 40 (34%) | 27 (27%) | 13 (81%) | 0.0004** |
| Plasma Cell Neoplasm | 42 (36%) | 41 (41%) | 1 (6%) | 0.06 |
| Hematologic Malignancy category, n (%) |  |  |  | 0.0002** |
| Lymphoid | 47 (41%) | 33 (33%) | 14 (88%) | 0.00005** |
| Myeloid | 27 (23%) | 26 (26%) | 1 (6%) | 0.11 |
| Plasma Cell Neoplasm | 42 (36%) | 41 (41%) | 1 (6%) | 0.02** |
| Prior COVID infection, n (%) |  |  |  |  |
| Yes | 15 (13%) | 13 (13%) | 2 (13%) | 1.00 |
| No | 101 (87%) | 87 (87%) | 14 (87%) |  |
| Cancer status at time vaccine, n (%) |  |  |  | 0.10 |
| Active | 49 (42%) | 46 (46%) | 3 (19%) | 0.17 |
| Progressive disease/relapse | 19 (17%) | 15 (15%) | 4 (25%) | 0.55 |
| Remission | 48 (41%) | 39 (39%) | 9 (56%) | 0.55 |
| Active cancer therapy at vaccination, n (%) |  |  |  |  |
| Yes | 69 (59%) | 60 (60%) | 9 (56%) | 0.79 |
| No | 47 (41%) | 40 (40%) | 7 (44%) |  |
| Transplant, n (%) |  |  |  | 0.79 |
| Autologous Stem Cell Transplant | 43 (37%) | 37 (37%) | 6 (38%) | 0.49 |
| Allogeneic Stem Cell Transplant | 4 (3%) | 3 (3%) | 1 (6%) | 0.49 |
| CAR-T therapy, n (%) | 7 (6%) | 1 (1%) | 6 (38%) | 0.00002** |
| Stem cell transplant, n (%) | 47 (41%) | 40 (40%) | 7 (44%) | 0.79 |
| Type of Vaccine, n (%) |  |  |  | 0.50 |
| Johnson & Johnson | 10 (9%) | 10 (10%) | 0 (0%) | 0.35 |
| Moderna | 36 (31%) | 30 (30%) | 6 (38%) | 0.57 |
| Pfizer | 70 (60%) | 60 (60%) | 10 (62%) | 1.0 |
| Last vaccine dose to Spike IgG testing (days), median [range] | 40 [14-154] | 45 [14-127] | 41 [15-154] | 0.71 |
| Spike IgG (AU/mL), median [range] | 2157 [<50-50000] | 3608 [51-50000] | < 50 | 0.32 |
| Johnson & Johnson | 1078 [138-50000] | 1078 [235-50000] | < 50 |  |
| Moderna | 5290 [50-50000] | 6766 [236-50000] | < 50 |  |
| Pfizer | 1697 [50-50000] | 2581 [51-50000] | < 50 |  |
| Adverse events post-first dose, n (%) |  |  |  | 0.71 |
| None | 64 (64%) | 55 (55%) | 9 (56%) | 0.77 |
| Mild/Moderate | 35 (35%) | 31 (31%) | 4 (25%) | 1.0 |
| Severe | 4 (4%) | 4 (4%) | 0 (0%) | 1.0 |
| Unknown | 13 (13%) | 10 (10%) | 3 (19%) | 0.39 |
| Adverse events post-second dose, n (%) | (N=106) | (N=90) | (N=16) | 0.92 |
| None | 53 (50%) | 44 (49%) | 9 (56%) | 0.77 |
| Mild/Moderate | 31 (29%) | 27 (30%) | 4 (25%) | 0.78 |
| Severe | 5 (5%) | 5 (5%) | 0 (0%) | 1.0 |
| Unknown | 16 (15%) | 14 (16%) | 3 (19%) | 0.72 |
| **Types of Cancer Therapy** |  |  |  |  |
| Antibody-drug conjugate, n (%) | 2 (2%) | 1 (1%) | 1 (6%) | 0.23 |
| Anti-CD19 antibody therapy, n (%) | 1 (1%) | 0 (0%) | 1 (6%) | 0.14 |
| Anti-CD20 antibody therapy, n (%) | 36 (31%) | 22 (22%) | 14 (88%) | 0.02** |
| Less than 6 months | 12 (10%) | 5 (5%) | 7 (44%) |  |
| Greater than 6 months | 24 (21%) | 17 (17%) | 7 (44%) |  |
| Anti-CD30 antibody therapy, n (%) | 1 (1%) | 1 (1%) | 0 (0%) | 1.00 |
| Anti-CD38 antibody therapy, n (%) | 16 (14%) | 15 (15%) | 1 (6%) | 0.69 |
| Anti-IL-6 antibody therapy, n (%) | 3 (3%) | 1 (1%) | 2 (13%) | 0.05 |
| Anti-viral therapy, n (%) | 4 (3%) | 3 (3%) | 1 (6%) | 0.45 |
| BCL-2 inhibitor, n (%) | 11 (9%) | 8 (8%) | 3 (19%) | 0.18 |
| BTK inhibitor, n (%) | 4 (3%) | 2 (2%) | 2 (13%) | 0.09 |
| CAR-T therapy, n (%) |  |  |  |  |
| Greater than 1 year | 5 (4%) | 0 (0%) | 5 (31%) | 0.29 |
| Spike IgG titer | 7 (6%) | 1 (1%) | 6 (38%) | 0.16 |
| Clinical Trial (experimental therapy), n (%) | 7 (6%) | 7 (7%) | 0 (0%) | 0.59 |
| Current Steroid Use, n (%) | 26 (22%) | 21 (21%) | 5 (31%) | 0.35 |
| Cytotoxic Chemotherapy, n (%) | 70 (60%) | 55 (55%) | 15 (94%) | 0.002** |
| Hormonal therapy (ADT, OFS, and AI), n (%) | 5 (4%) | 5 (5%) | 0 (0%) | 1.00 |
| Immune checkpoint inhibitor, n (%) | 2 (2%) | 1 (1%) | 1 (6%) | 0.26 |
| Immunomodulatory agent, n (%) | 40 (34%) | 39 (39%) | 1 (6%) | 0.01** |
| IVIG, n (%) | 8 (7%) | 4 (4%) | 4 (25%) | 0.01** |
| No treatment, n (%) | 5 (4%) | 5 (5%) | 0 (0%) | 1.00 |
| Proteasome inhibitor, n (%) | 39 (34%) | 38 (38%) | 1 (6%) | 0.01** |
| Radiation, n (%) | 22 (19%) | 20 (20%) | 2 (13%) | 0.73 |
| Stem cell transplant, n (%) |  |  |  |  |
| Greater than 1 year | 38 (33%) | 32 (32%) | 6 (38%) | 0.78 |
| Spike IgG titer | 47 (41%) | 40 (40%) | 7 (44%) | 0.69 |
| Supportive Care, n (%) | 11 (9%) | 11 (11%) | 0 (0%) | 0.36 |
| Surgery, n (%) | 11 (9%) | 10 (10%) | 1 (6%) | 1.00 |
| TGFβ inhibitor, n (%) | 4 (3%) | 4 (4%) | 0 (0%) | 1.00 |
| Tyrosine kinase inhibitor, n (%) | 5 (4%) | 5 (5%) | 0 (0%) | 1.00 |

ADT, Androgen deprivation therapy; BCL-2, B cell lymphoma 2; BTK, Bruton's tyrosine kinase; OFS, Ovarian function suppression; AI, Aromatase inhibitors; IVIG, Intravenous immune globulin; TGFβ, Transforming growth factor beta.
\*Acute leukemia (ALL, Acute lymphocytic leukemia; AML, Acute myeloid leukemia); MDS, Myelodysplastic syndrome; MPN, Myeloproliferative neoplasm (CML, Chronic Myeloid Leukemia; MDS/MPN); Non-Hodgkin Lymphoma (ATLL, Adult T-cell Leukemia/Lymphoma; Burkitt's Lymphoma; CLL, Chronic Lymphocytic Lymphoma; DLBCL, Diffuse Large B-cell Lymphoma; Follicular Lymphoma; MALT Lymphoma; Mantle Cell Lymphoma; Marginal Zone Lymphoma; Mycosis Fungoides; Peripheral T-cell Lymphoma; Primary CNS Lymphoma; Waldenstrom's Macroglobulinemia); Plasma Cell Neoplasm (Multiple Myeloma; Amyloidosis; Plasma Cell Leukemia)
\*\* = Statistically significant

Collectively, our data mirrors the current literature showing significantly lower rates of seroconversion in recipients following highly immunosuppressive therapies such as anti-CD20 therapies, and CAR-T therapy; however, we also noted that patients simply on cytotoxic chemotherapy are also at risk of poor seroconversion. Similar to the recent prospective cohort registry study through the Leukemia and Lymphoma Society,^14^ we have noted that patients with myeloid malignancies and plasma cell neoplasms do quite well with COVID-19 vaccination despite their treatment history, with immunomodulatory agents and proteosome inhibitors actually correlating with higher seroconversion rates. We also observed an 86% seroconversion rate in autologous transplant recipients, higher than the previously reported 66%.^12^ Recent anti-CD20 therapy (less than 6 months) and any time post CD19 CAR-T therapy predicts high rates of seronegativity showing that these therapies suppress normal B cell function for long durations after dosing.^15,16^ It may be important to vaccinate all CAR-T therapy patients prior to lymphodepleting chemotherapy and consider treatment interruption for those patients on anti-CD20 therapy when feasible. In addition, despite our concordant findings with the current literature, race has not been a topic of discussion for successful seroconversion after vaccination. It is well known that race is an important contributor to morbidity and mortality in patients with COVID-19,^17^ especially in the hematologic malignancy population, with non-White patients having a significantly higher risk of mortality than White patients,^1^ and a higher rate of vaccine hesitancy.^18^ Most studies described thus far do not stratify by race with those that do only ranging between 3.5-20% non-White patients.

To our knowledge, this is the first report on high seroconversion rates after SARS-CoV-2 vaccination in non-White patients with hematologic malignancies treated with a wide spectrum of therapeutic modalities. Vaccination is safe, effective, and should be encouraged in patients with hematologic malignancies. Our minorities based study could be employed as an educational tool to dispel myths and provide data driven evidence to overcome vaccine hesitancy.

## Data Availability

Please contact Dr. Alejandro Sica regarding availability of data.

## Acknowledgments

We acknowledge Albert Einstein Cancer Center grant P30 CA013330 and NCORP grant 2UG1CA189859-06 in providing funding for this project. This work was supported partly by the Jane A. and Myles P. Dempsey fund.

## Authorship Contributions

LCS, AT, BH, AV, LB-R, and AS designed the study. LCS, RG, AT, JDG-L, AHB, SR, KF, MA, NK, KG, MG, AS, IM, IB, BH, AV, MC, LBR and AS participated in patient recruitment and data collection. KP and LCS analyzed and interpreted the data. LCS wrote the first draft of manuscript and AT and AS provided a critical review of the letter’s content. All authors reviewed the manuscript and approved the final letter.

Correspondence: Dr. R. Alejandro Sica, Department of Oncology, Montefiore Medical Center/Albert Einstein College of Medicine, 111 E 210^th^ St, Bronx, NY, 10467.

